# Social networks and their association with quality of life among older adults in rural Burkina Faso

**DOI:** 10.64898/2026.06.21.26356165

**Authors:** Indrakantha Welgama, Dina Goodman-Palmer, Guy Harling, Sandra Agyapong-Badu, Carolyn Greig, Maxime Inghels, Miles D Witham, Lucienne Ouermi, Boubacar Coulibaly, Lisa R Hirschhorn, Mamadou Bountogo, Ali Sie, Justine Davies

## Abstract

**Objective:** This study aimed to identify the types of social networks present among older adults in a rural, low-income country setting and describe their association with quality of life (QoL).

**Methods:** A population-representative, cross-sectional survey was conducted in 60 villages around Nouna in Burkina Faso from July to August 2021. Data were collected from resident adults aged 40 years and older. Variables captured were sociodemographic status; social network characteristics (using the Practitioner Assessment of Network Typology (PANT)); quality of life (using the EuroHIS-8 tool); presence of non-communicable diseases, mental health conditions, and disability. Additionally, social networks were broadly categorised as aggregated integrated and aggregated less-integrated groups. Social network types and the groups were described separately, and a multivariable linear regression model was used to understand the association between social network types and QoL, adjusted for sociodemographic and morbidity factors.

**Results:** Among the 2390 respondents, median age was 55 yrs (IQR: 47-64 yrs) and 55.8% were female. Locally Integrated (35.4%) or Family Dependent (30.3%) were the most common PANT social network types, followed by a mixed group (having characteristics of two or more social network types) (30.5%). Private Restricted (2.1%), Locally Self-Contained (1.2%), and Wider Community-Focussed (0.4%) types were uncommon. Adults with aggregated integrated network groups (36.1%) and aggregated less-integrated group (36.0%) were near equal, while others were non-aggregable. Although Wider Community-Focused type showed a significantly better QoL (β= 8.69, 95%CI: 4.10 to 13.27), the association between social networks and QoL were subdued when controlled for morbidity factors, and hence no significant associations were observed between other types or the aggregated groups.

**Conclusion:** Although having integrated social networks lead to a better QoL, morbidity has a greater effect on the QoL among older adults in Nouna and hence, investing more on improving the physical and mental health needs appears more beneficial.

## Introduction

Social support networks, also referred to as social networks, are the constellation of relationships within which a person interacts in their day to day life [1]. They are an important source of personal, mental and social support across all stages of life [2–4]. Such networks may include immediate and close family members, friends, neighbours, colleagues, and wider community groups or individuals [2, 5, 6]. Social networks themselves are affected by the traits of individuals (their demographic, personality, and socio-economic characteristics), the community and socio-cultural norms in which people live, and the family structures and expectations in which people grow up [7, 8]. Although certain studies have categorised social networks based on criteria relevant to their studies [9, 10], the Practitioner Assessment of Network Typology (PANT) developed by Wenger has been frequently used in both high- and middle-income countries [2, 7, 8, 11–13]. This categorises social networks of older persons as Family Dependent (mostly reliant on family for social contact and support); Locally Integrated (with networks formed from both family interactions and those within the local community); Locally Self-Contained (with some, although limited, interactions with both family and community); Wider Community-Focussed (with most interactions with the community and less with family); and Private Restricted (with few interactions with family or community). In addition, Wenger has also indicated that about 20% of the community would have characteristics of multiple network types [11], and they are referred to as inconclusive or borderline group by prior studies [2, 13, 14]. Distribution of social networks and their relationship with outcomes such as health and QoL, are contextually determined. However, studies suggest that across contexts, the Locally Integrated network is the most common, and has the strongest association with beneficial outcomes [5, 8].

Quality of Life (QoL) is defined as “an individual’s perception of their position in life in the context of the culture and value systems in which they live, and in relation to their goals, expectations, standards and concerns” [15]. As well as being important in its own right, QoL is recognised to be strongly associated with an increased hazard of mortality in the short, medium, or long term [16]. Given increasing number of people living into older age in low- or middle-income countries, it is important to ensure that good QoL is attained and maintained into older ages. In addition to socio-economic, demographic, and health factors, social support networks have been shown to be important determinants of QoL in older people [9, 15, 17].

Most information on social network types and their association with QoL comes from high income countries, with some from middle income countries [4, 6, 8, 18]. There is little knowledge from low-income countries (LICs) on the social network types of older adults or their associations with QoL. Considering the numerous adverse socio-cultural and financial challenges encountered by the people in LICs which can affect the formation of social networks and in turn, the QoL, it cannot be assumed that evidence from high- or middle-income countries will translate to LICs [10, 19, 20]. Therefore, this study aimed to describe the social network types of older adults in a LIC setting, factors associated with their social network types, and the associations between these social networks and the QoL.

## Methods

### Study setting

Burkina Faso is a landlocked, low-income country in the Western Sub-Saharan Africa. It is one of the least developed countries in the world with a per capita GNI of US$ 840 in 2021 [21]. This study was conducted in the Nouna Health and Demographic Surveillance System (HDSS) site in the Boucle du Mouhoun province of the north-western region of Burkina Faso [22]. The HDSS site is situated in an area of 1758km^2^ of dry savannah land, comprising of Nouna town and 59 surrounding rural villages [22]. The main socioeconomic activity in the region is subsistence farming and animal husbandry [22, 23].

### Data collection

Data were collected as part of the “Centre de Recherche en Santé de Nouna (CRSN) Heidelberg Aging Study” (CHAS) population representative, cross sectional survey [24]. The CHAS data were collected between 1^st^ July 2021 and 22^nd^ August 2021. A survey was administered by trained data collectors and responses were collected electronically using Open-Data-Kit software [25]. The survey was developed in French (the national language of Burkina Faso) and translated into local, oral languages during administration. Data collectors were multilingual, and real-time translations formed part of their training.

### Study population

The inclusion criteria of the sample were: being aged 40 years or older, resident in a HDSS household for at least the past six months and providing written informed consent to participate in the study. We selected the age threshold of 40 years given that life expectancy in Burkina Faso at the time of this study was only 62.3 years, and under 5% of the national population were aged 60 years or older [26, 27]. Prior research in this population shows that the biological age of people over 40 years is similar to those in high income countries who are two decades older [24, 28, 29]. No specific exclusion criteria were considered for this study. In each HDSS village, 50 houses were randomly selected from all households containing one or more eligible adults over 40 years of age and from each selected house, one eligible adult was randomly selected. Alternatively, in villages where the eligible population was less than 50, all eligible adults in households were included in the sample. Written informed consent was obtained from all selected participants; those who were illiterate were assisted by a literate representative who was not a member of the study team [24, 30].

### Variables included

To inform variables to include in our analysis, we reviewed literature on social networks and QoL to develop a conceptual framework (supplementary material 1) of the factors which could influence the relationship between social networks and QoL among older adults in Nouna. Resulting socio-demographic variables were age (continuous), sex (male or female), and marital status (living alone [single, divorced or widowed] or living with partner [either married or cohabiting]). Religion was categorized into three groups: the two most common (Islam or Christianity [including Catholics and Protestants]) separately and all other religions (including Animists and other traditional religions) together as one group. Ethnicity was categorised into three groups: the two most common (Bwaba or Dafin) separately and all other ethnicities (including Mossi, Bobo, Dioula, Guransi, Fulani, Lobi, Tureg and other minor ethnic groups) as a single group. Education status (no formal education versus having any education) and another 37 questions related to household assets and dwelling characteristics were also collected. Wealth quintiles were derived from the information related to household assets using the method described by Filmer and Pritchet [31].

The presence of non-communicable diseases (NCD) morbidity was determined based on self-reporting on a prior diagnosis of at least one of the following conditions: high blood pressure, diabetes, heart disease, hypercholesterolaemia, chronic respiratory disease, stroke or cancer [29]. Symptoms of depression were assessed using Patient Health Questionnaire-9 (PHQ-9) [32], and categorized as mild or no symptoms of depression in those with a PHQ-9 score of less than 10, or moderate to severe depression in those with a score of greater than or equal to 10 [32]. Symptoms of anxiety were assessed using the Generalized Anxiety Disorder 2-item (GAD-2) questionnaire [33], and respondents with a GAD-2 score of greater than or equal to 3 were classified as having symptoms of anxiety [34]. Symptoms of dementia were assessed using the Community Screening Instrument for Dementia (CSI-D) [35], and the possibility of having dementia was defined as having a CSI-D score of less than 7 out of 10 [35, 36]. The presence of symptoms of at least one of anxiety, depression, or dementia was considered as the presence of mental health morbidity. Disability status was assessed using the 12-item WHO Disability Assessment Schedule - version 2 (WHODAS V2.0) [37]. WHODAS scores were normalised to a continuous data scale of 0–100, where 0 indicated no disability and 100 represented the maximum disability [29, 37]. Quality of Life (QoL) was assessed using the EuroHIS-8, an 8-item version of the WHO Quality of Life (WHOQoL) questionnaire [38]. EuroHIS-8 scores were normalised to a continuous data scale of 0-100, with higher values representing better QoL [29, 38].

We utilised Wenger’s eight-item PANT questions to determine the respondent’s social support network type, with questions exploring the accessibility and contact frequency of family, friends, neighbours and community groups [7]. If a respondent had characteristics suggestive of having two or more social network types, they were categorised into a mixed type. We also aggregated participants into two groups – an “Aggregate integrated group” (containing Locally Integrated, Wider Community-Focussed, and people in the mixed Group with a mixture of characteristics of these two groups) and an “Aggregate less-integrated group” (containing Family Dependent, Locally Self-contained, and Private Restricted types and people in the mixed group with a mixture of characteristics of these types). Further details related to the categorisation of PANT social networks are in supplementary material 2.

### Statistical analysis

Data were analysed using STATA 18.0 (StataCorp, USA) [39]. Categorical variables were presented as count and percentage; continuous variables were presented as mean and standard deviation, or median and inter-quartile range (IQR), depending on their distribution. Variables were described in total and disaggregated by sex. We used weights generated using the inverse probability of response for survey sample participation, calculated using a sex-specific logistic model from the sample including age, religion, ethnicity and village of residence.

To evaluate associations between social network types and sociodemographic and morbidity characteristics, a weighted multinomial logistic regression model was developed with social network types as the dependent variable, using the most common social network type as the reference category. Relative risk ratios (RRR) with 95% confidence intervals were calculated to report outcomes.

To describe the association between QoL and social network types, we conducted sequential multivariable analysis using linear regression with the normalised EuroHIS-8 score as the dependent variable. We first included the social network types in the model (model 1), then added sociodemographic factors (model 2), and finally, added the morbidity and disability factors (model 3). Results were reported as the regression coefficient (β) with 95% confidence intervals.

Our main analyses excluded the mixed group from the model due to the presence of multiple different social network characteristics in this group. In an exploratory analysis, we repeated the linear regression of the relationship between QoL and social network types in the aggregate groups.

We generated marginal means plots to observe the estimated QoL scores for each of the PANT social networks (model 1), and when controlling for the sociodemographic (model 2) and morbidity factors (model 3). Similar marginal means plots were generated to understand the relationship between the QoL and the aggregated social network groups.

## Results

A total of 4100 eligible participants were approached of whom 3089 consented to be enrolled in the study. Of them, 2390 (77.4%) responded to the PANT questions and were included in the analysis. Among the respondents, age ranged from 40 to 105 years, with a median of 55 years (IQR 47 to 64). The majority (1334, 55.8%) were females. At least one NCD morbidity was observed in 509 (21.3%) participants; 350 (14.6%) participants had symptoms indicative of mental health morbidity; median disability score was 18.7 (IQR: 18.7 to 25.0), and median quality of life score was 59.4 (IQR: 50.0 to 65.6). (Table 1).

**Table 1.**
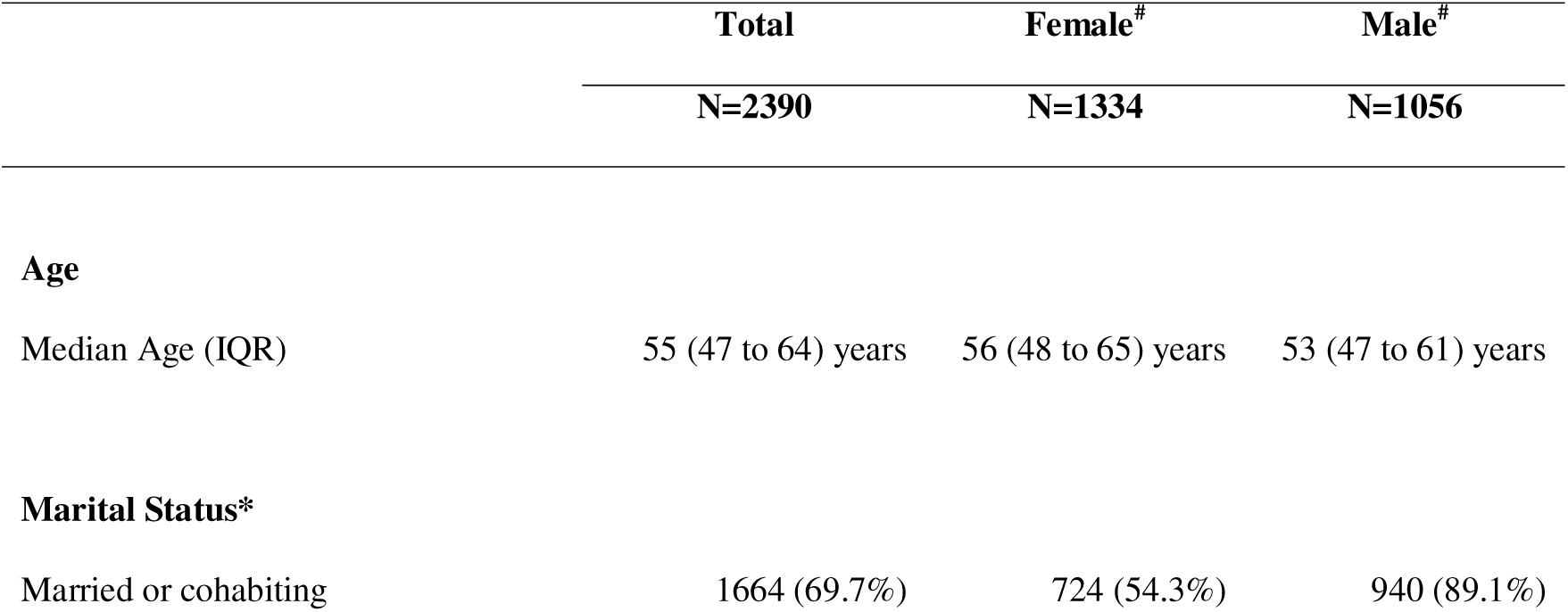
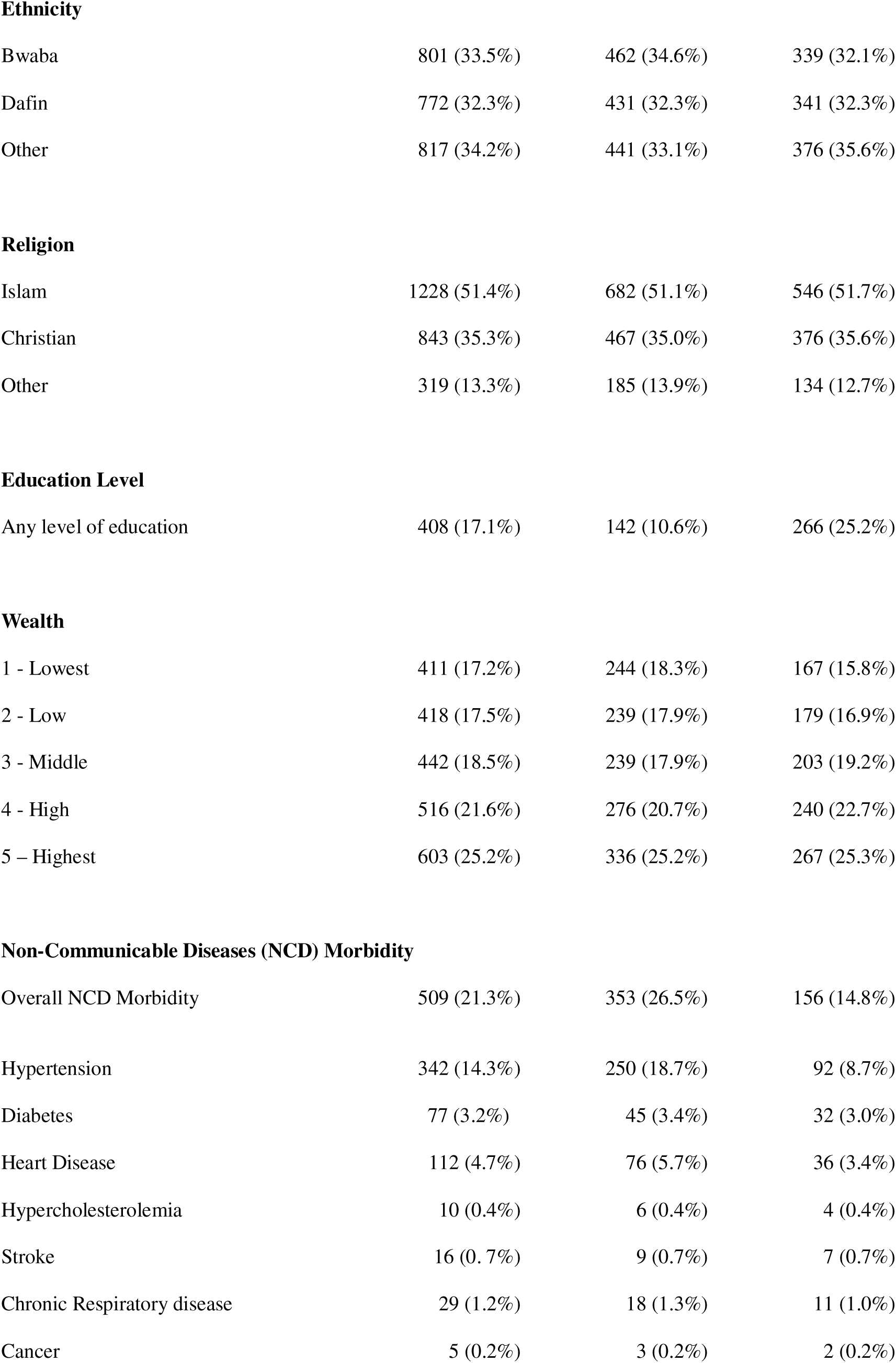
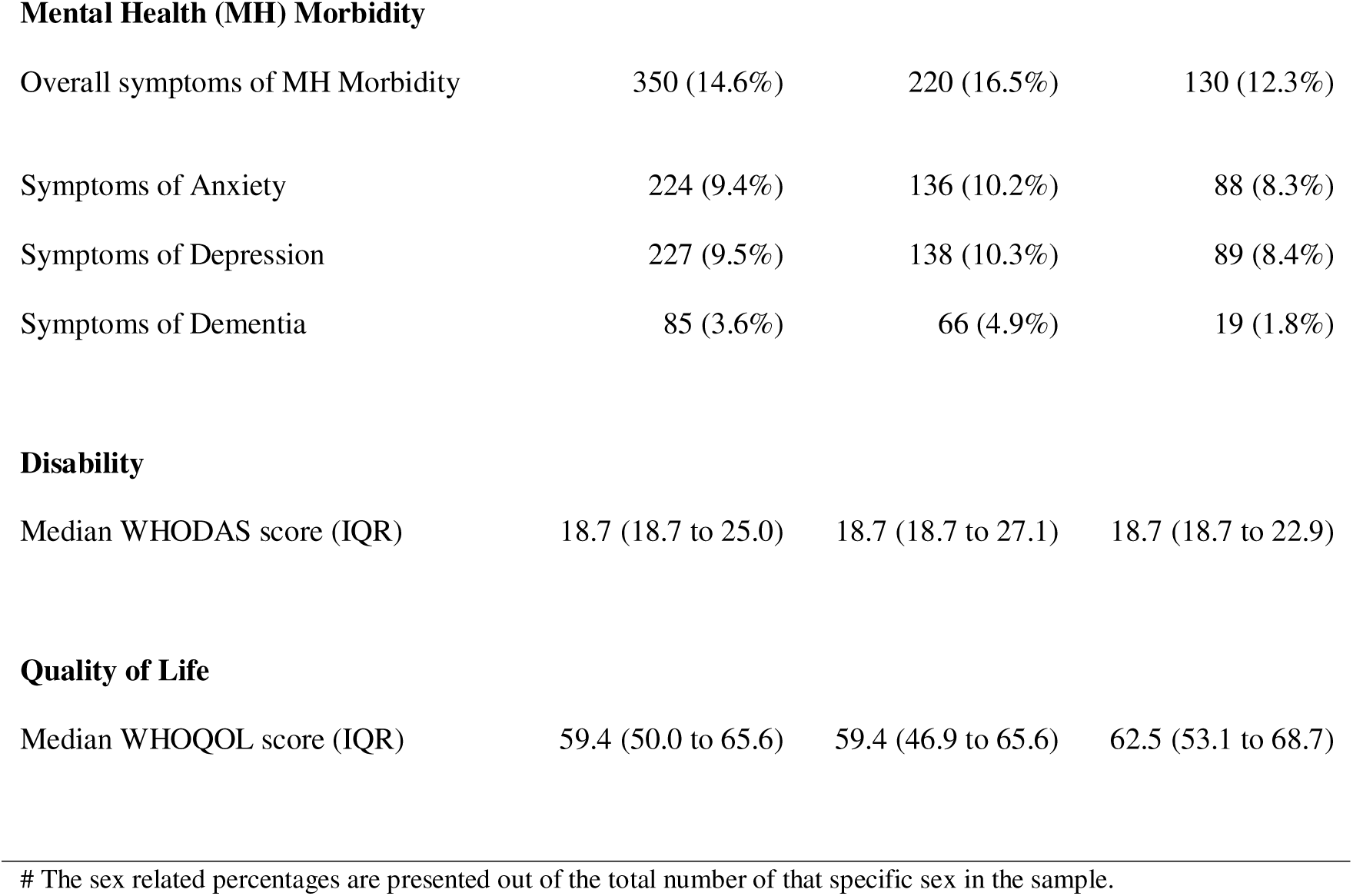
Sociodemographic and morbidity characteristics (total and sex-stratified count and percentage, or the median and IQR where indicated)

|  | **Total** | **Female<sup>#</sup>** | **Male<sup>#</sup>** |
| --- | --- | --- | --- |
|  | **N=2390** | **N=1334** | **N=1056** |
| **Age** |  |  |  |
| Median Age (IQR) | 55 (47 to 64) years | 56 (48 to 65) years | 53 (47 to 61) years |
| **Marital Status\*** |  |  |  |
| Married or cohabiting | 1664 (69.7%) | 724 (54.3%) | 940 (89.1%) |
| **Ethnicity** |  |  |  |
| Bwaba | 801 (33.5%) | 462 (34.6%) | 339 (32.1%) |
| Dafin | 772 (32.3%) | 431 (32.3%) | 341 (32.3%) |
| Other | 817 (34.2%) | 441 (33.1%) | 376 (35.6%) |
| **Religion** |  |  |  |
| Islam | 1228 (51.4%) | 682 (51.1%) | 546 (51.7%) |
| Christian | 843 (35.3%) | 467 (35.0%) | 376 (35.6%) |
| Other | 319 (13.3%) | 185 (13.9%) | 134 (12.7%) |
| **Education Level** |  |  |  |
| Any level of education | 408 (17.1%) | 142 (10.6%) | 266 (25.2%) |
| **Wealth** |  |  |  |
| 1 - Lowest | 411 (17.2%) | 244 (18.3%) | 167 (15.8%) |
| 2 - Low | 418 (17.5%) | 239 (17.9%) | 179 (16.9%) |
| 3 - Middle | 442 (18.5%) | 239 (17.9%) | 203 (19.2%) |
| 4 - High | 516 (21.6%) | 276 (20.7%) | 240 (22.7%) |
| 5 – Highest | 603 (25.2%) | 336 (25.2%) | 267 (25.3%) |
| **Non-Communicable Diseases (NCD) Morbidity** |  |  |  |
| Overall NCD Morbidity | 509 (21.3%) | 353 (26.5%) | 156 (14.8%) |
| Hypertension | 342 (14.3%) | 250 (18.7%) | 92 (8.7%) |
| Diabetes | 77 (3.2%) | 45 (3.4%) | 32 (3.0%) |
| Heart Disease | 112 (4.7%) | 76 (5.7%) | 36 (3.4%) |
| Hypercholesterolemia | 10 (0.4%) | 6 (0.4%) | 4 (0.4%) |
| Stroke | 16 (0.7%) | 9 (0.7%) | 7 (0.7%) |
| Chronic Respiratory disease | 29 (1.2%) | 18 (1.3%) | 11 (1.0%) |
| Cancer | 5 (0.2%) | 3 (0.2%) | 2 (0.2%) |
| **Mental Health (MH) Morbidity** |  |  |  |
| Overall symptoms of MH Morbidity | 350 (14.6%) | 220 (16.5%) | 130 (12.3%) |
| Symptoms of Anxiety | 224 (9.4%) | 136 (10.2%) | 88 (8.3%) |
| Symptoms of Depression | 227 (9.5%) | 138 (10.3%) | 89 (8.4%) |
| Symptoms of Dementia | 85 (3.6%) | 66 (4.9%) | 19 (1.8%) |
| **Disability** |  |  |  |
| Median WHODAS score (IQR) | 18.7 (18.7 to 25.0) | 18.7 (18.7 to 27.1) | 18.7 (18.7 to 22.9) |
| **Quality of Life** |  |  |  |
| Median WHOQOL score (IQR) | 59.4 (50.0 to 65.6) | 59.4 (46.9 to 65.6) | 62.5 (53.1 to 68.7) |

A clear PANT social network type was identifiable among 1660 (69.5%) participants while the remaining 730 (30.5%) with characteristics of two or more types were included as a mixed group. The most common social network types were the Locally Integrated (35.4%) followed by Family Dependent (30.3%) (Table 2). An in-depth exploration of the mixed group is presented in supplementary material 3. Considering the aggregated groups, the Aggregated integrated group had 862 (36.1%) participants, with a near similar number in the Aggregated less-integrated group (861, 36.0%) (Table 2). The sociodemographic and morbidity characteristics of individuals having each of these PANT social network types are also presented in supplementary material 4.

**Table 2.**
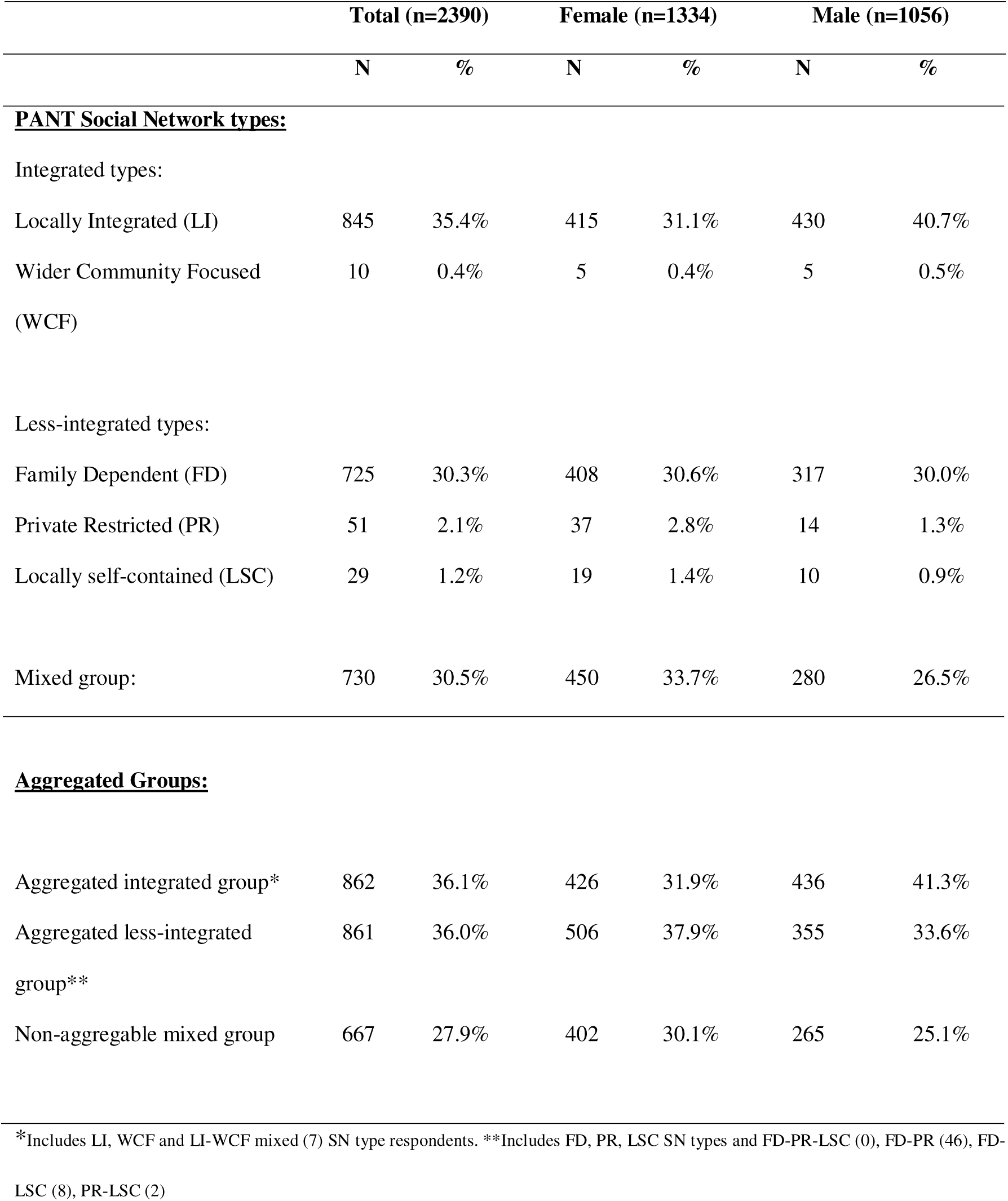
Total and sex-disaggregated distribution of PANT social network types and the Aggregated social network groups.

In the multinomial logistic regression model of participants having specific PANT social network types, the Locally Integrated social network type was considered as the reference (Table 3). In comparison, older age was associated with a higher possibility of having a Private Restricted (RRR=1.08, p<0.01) or Locally Self-contained network type (RRR=1.06, p=0.01), while a relatively younger age was associated with having a Wider Community-Focussed social network type (RRR=0.92, p=0.03). Men were less likely to have a Private Restricted network type (RRR = 0.21, p=0.01) than women. Participants with mental health morbidity were more likely to have a Family Dependent network type (RRR=1.67, p=0.01). No significant associations were observed between social network types and level of education, wealth quintiles, NCD morbidity, or disability (Table 3). Including the mixed group in the model did not substantially affect the results (supplementary material 5).

**Table 3.** Weighted multinomial logistic regression model of PANT social network types and their association with the sociodemographic and morbidity factors, in reference to Locally Integrated network type (n=1660)

| Characteristic | PANT Social Network types |  |  |  |  |  |  |  |  |  |  |  |
| --- | --- | --- | --- | --- | --- | --- | --- | --- | --- | --- | --- | --- |
|  | Family Dependent |  |  | Private Restricted |  |  | Locally Self-Contained |  |  | Wider Community-Focused |  |  |
|  | RRR | 95% CI | p-value | RRR | 95% CI | p-value | RRR | 95% CI | p-value | RRR | 95% CI | p-value |
| **Age** |  |  |  |  |  |  |  |  |  |  |  |  |
| Age in years | 0.99 | [0.98 to 1.01] | 0.65 | **1.08** | [1.03 to 1.14] | <0.01 | **1.06** | [1.01 to 1.12] | 0.01 | **0.92** | [0.86 to 0.99] | 0.03 |
| **Sex** |  |  |  |  |  |  |  |  |  |  |  |  |
| Female | Ref |  |  | Ref |  |  | Ref |  |  | Ref |  |  |
| Male | 0.78 | [0.57 to 1.06] | 0.11 | **0.21** | [0.06 to 0.71] | 0.01 | 0.47 | [0.14 to 1.64] | 0.24 | 1.28 | [0.16 to 10.2] | 0.81 |
| **Marital Status** |  |  |  |  |  |  |  |  |  |  |  |  |
| Single/ divorced/<br>widowed | Ref |  |  | Ref |  |  | Ref |  |  | Ref |  |  |
| Married /<br>cohabiting | 0.85 | [0.60 to 1.22] | 0.38 | 0.89 | [0.30 to 2.65] | 0.83 | 0.45 | [0.15 to 1.37] | 0.16 | 0.98 | [0.03 to 31.9] | 0.99 |
| **Ethnicity** |  |  |  |  |  |  |  |  |  |  |  |  |
| Bwaba | Ref |  |  | Ref |  |  | Ref |  |  | Ref |  |  |
| Dafin | 0.93 | [0.61 to 1.43] | 0.75 | 0.45 | [0.11 to 1.79] | 0.26 | 0.65 | [0.15 to 2.83] | 0.57 | 6.07 | [0.55 to 66.9] | 0.14 |
| Other | 1.03 | [0.65 to 1.63] | 0.90 | 2.45 | [0.73 to 8.26] | 0.15 | 1.32 | [0.33 to 5.34] | 0.69 | 3.85 | [0.18 to 82.1] | 0.39 |
| **Religion** |  |  |  |  |  |  |  |  |  |  |  |  |
| Islam | Ref |  |  | Ref |  |  | Ref |  |  | Ref |  |  |
| Christian | **0.60** | [0.41 to 0.88] | 0.01 | 0.65 | [0.21 to 2.00] | 0.45 | 0.94 | [0.27 to 3.28] | 0.93 | 1.04 | [0.13 to 8.04] | 0.98 |
| Other | 0.58 | [0.34 to 1.00] | 0.05 | 0.22 | [0.04 to 1.26] | 0.09 | 1.08 | [0.23 to 5.10] | 0.92 | 1.82 | [0.24 to 14.0] | 0.56 |
| **Education Level** |  |  |  |  |  |  |  |  |  |  |  |  |
| No Formal Schooling | Ref |  |  | Ref |  |  | Ref |  |  | Ref |  |  |
| Any level of education | 1.64 | [0.37 to 1.54] | 0.85 | 1.64 | [0.37 to 7.28] | 0.51 | 2.45 | [0.78 to 7.71] | 0.12 | 1.39 | [0.40 to 4.89] | 0.61 |
| **Wealth Quintiles** |  |  |  |  |  |  |  |  |  |  |  |  |
| 1. Lowest | Ref |  |  | Ref |  |  | Ref |  |  | Ref |  |  |
| 2. Low | 0.89 | [0.53 to 1.37] | 0.50 | 0.27 | [0.07 to 1.05] | 0.06 | 2.31 | [0.44 to 11.9] | 0.32 | NR | NR | NR |
| 3. Middle | 1.48 | [0.94 to 2.36] | 0.09 | 0.66 | [0.23 to 1.90] | 0.44 | 2.19 | [0.33 to 14.5] | 0.41 | NR | NR | NR |
| 4. High | 1.47 | [0.93 to 2.33] | 0.10 | 0.25 | [0.05 to 1.26] | 0.09 | **5.32** | [1.08 to 26.2] | 0.04 | 0.65 | [0.08 to 5.37] | 0.69 |
| 5. Highest | 1.51 | [0.95 to 2.39] | 0.08 | 0.38 | [0.12 to 1.18] | 0.09 | 2.38 | [0.43 to 13.2] | 0.32 | 0.67 | [0.08 to 5.83] | 0.72 |
| **NCD Morbidity** |  |  |  |  |  |  |  |  |  |  |  |  |
| Absent | Ref |  |  | Ref |  |  | Ref |  |  | Ref |  |  |
| Present | 1.28 | [0.92 to 1.80] | 0.15 | 1.18 | [0.51 to 2.71] | 0.70 | 0.72 | [0.24 to 2.21] | 0.57 | 1.83 | [0.32 to 10.6] | 0.49 |
| **MH Morbidity** |  |  |  |  |  |  |  |  |  |  |  |  |
| Absent | Ref |  |  | Ref |  |  | Ref |  |  | Ref |  |  |
| Present | **1.67** | [1.11 to 2.50] | 0.01 | 2.19 | [0.74 to 6.42] | 0.15 | 2.17 | [0.85 to 5.54] | 0.11 | NR | NR | NR |
| **Disability** |  |  |  |  |  |  |  |  |  |  |  |  |
| WHODAS Score | 1.01 | [0.99 to 1.02] | 0.07 | 1.02 | [0.99 to 1.05] | 0.11 | 1.00 | [0.97 to 1.03] | 0.77 | 0.99 | [0.93 to 1.06] | 0.94 |

RRR - Relative Risk ratio, 95% CI - 95% Confidence Interval, p-value - Probability value, Ref – Reference variable class, NR – Not reported. Significant associations are depicted with a bold font RRR. Note: Mixed group has not been included.

For the sequential linear regression exploring associations between the types of PANT social networks and QoL, the Locally Integrated social network type was the reference (Table 4). Among the PANT social network type, those with a Wider Community-Focussed network had a significantly better QoL (β = 11.88, p < 0.01) which remained same when controlled for sociodemographic factors (in models 2) and morbidity factors (in model 3). The association between a worse QoL and being in the Family Dependent (β = −2.53, p < 0.01) or the Private Restricted network types (β = −5.71, p =0.04) in model 1, was reduced in models 2 and 3 when controlling for sociodemographic and morbidity factors. No association was observed between the Locally Self-Contained type and QoL. Younger age, being a male, or belonging to a higher wealth quintile, absence of NCD morbidity, and lower disability were associated with a better QoL. Including the Mixed type didn’t result in any substantial difference in the associations (supplementary material 6). Estimated marginal means plots are shown in Figure 1.

**Figure 1.**
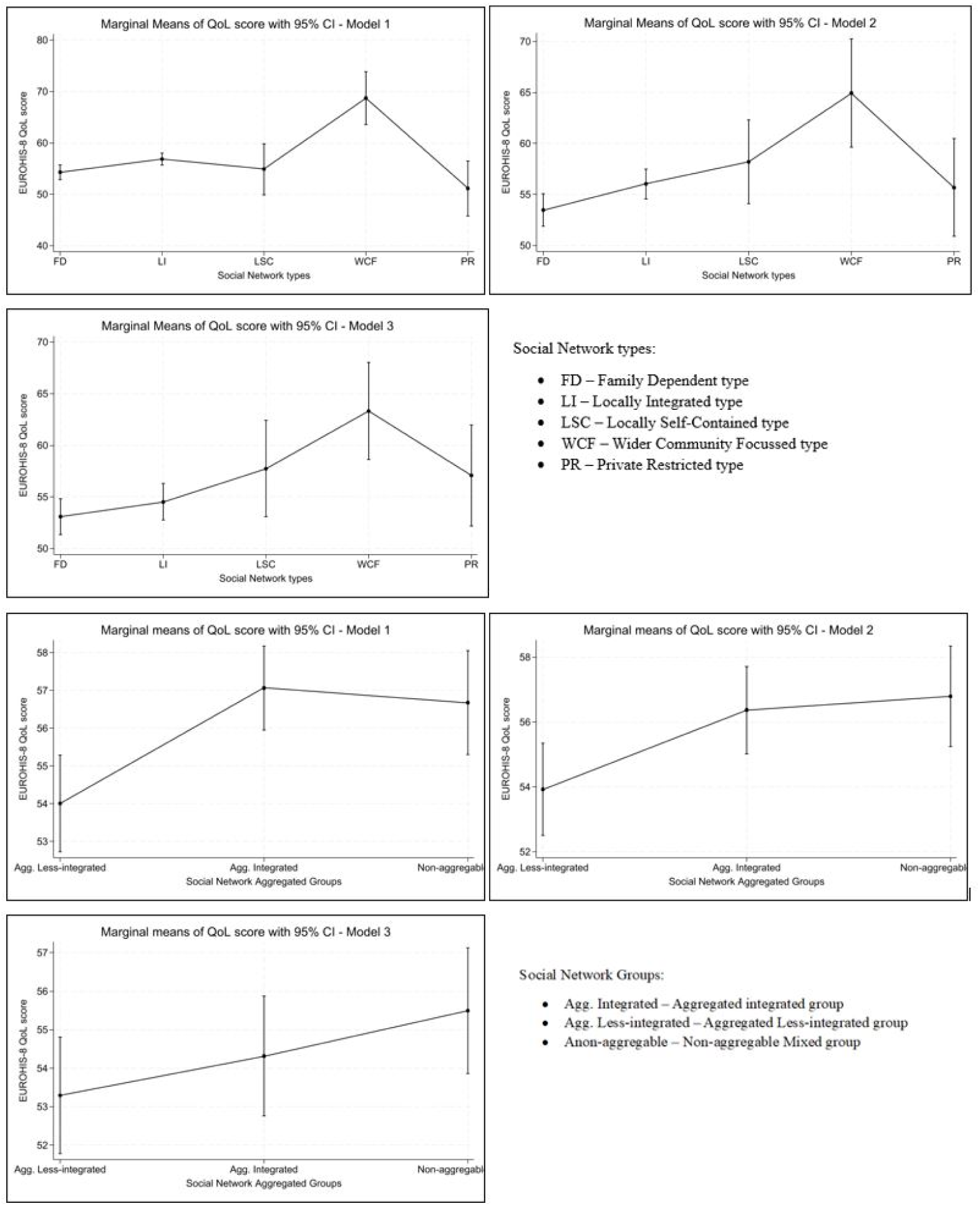
Estimated marginal means and the 95% confidence intervals for the normalised EUROHIS-8 Quality of Life scores for PANT social network types, and for aggregated social network groups.

**Table 4.**
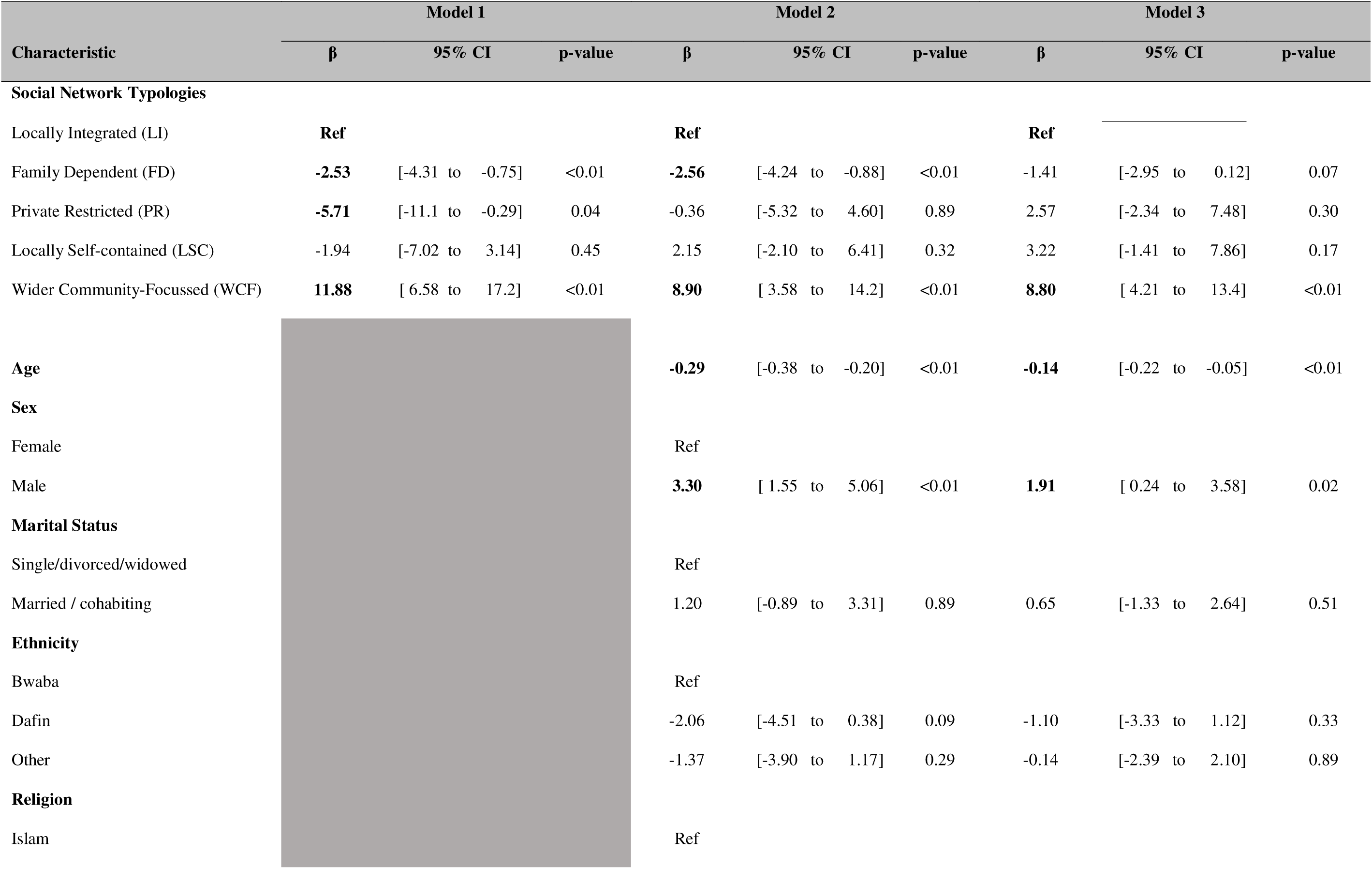
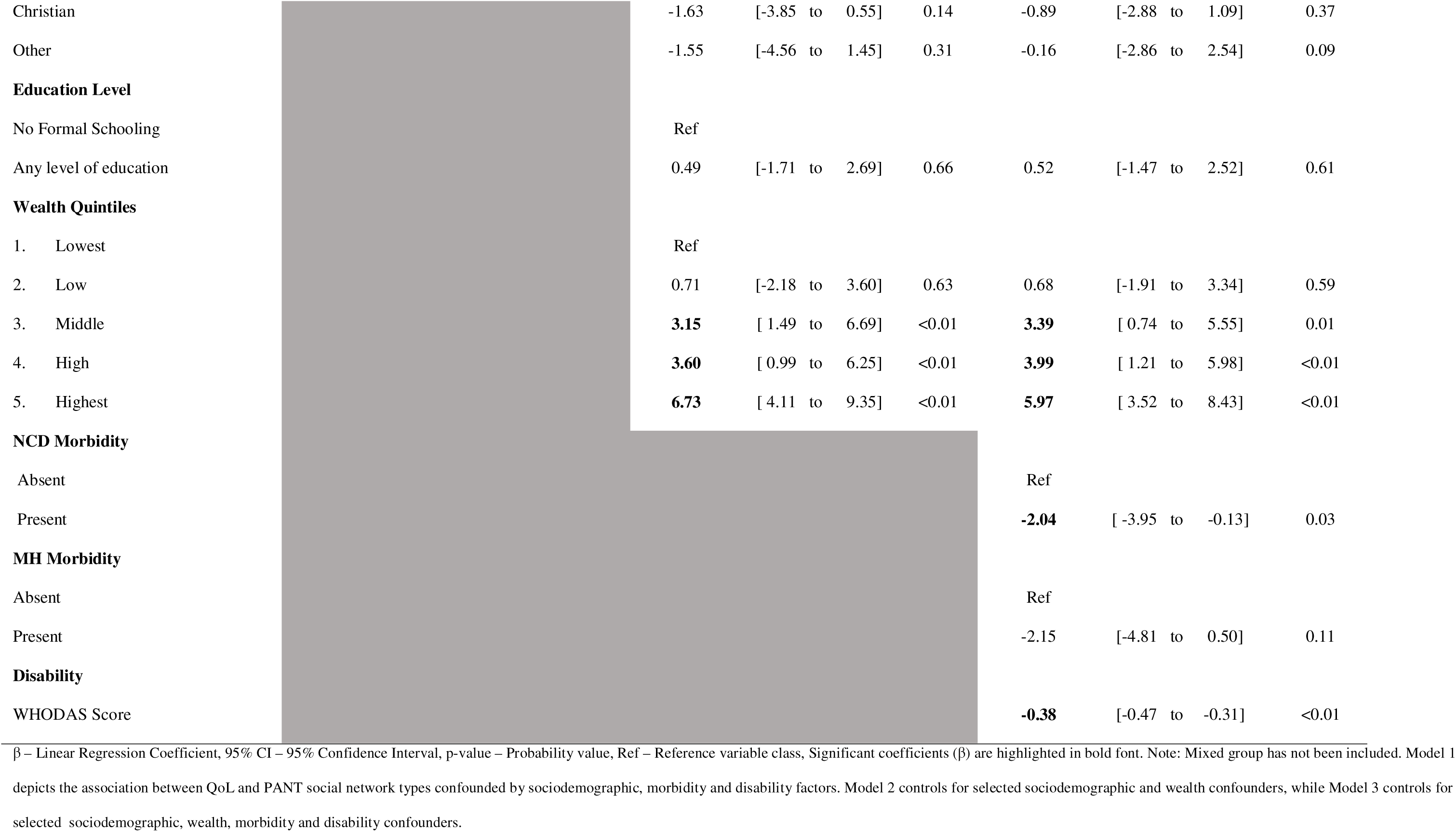
Weighted linear regression models for relationship between WHO quality of life score and PANT social network types, in the presence of sociodemographic and morbidity factors (n=1660)

The associations between QoL and aggregate network groups (Table 5) showed that being in the aggregated less-integrated group was associated with a significantly worse QoL in model 1 and 2, but this association disappeared in model 3 when controlling for morbidity covariables. The significantly lower QoL among females also disappeared when morbidity factors were added (model 3). The associations between the aggregated social network groups and other sociodemographic and morbidity factors remained similar to the findings of the analysis using PANT network types. Estimated marginal mean plots are shown in Figure 1.

**Table 5.**
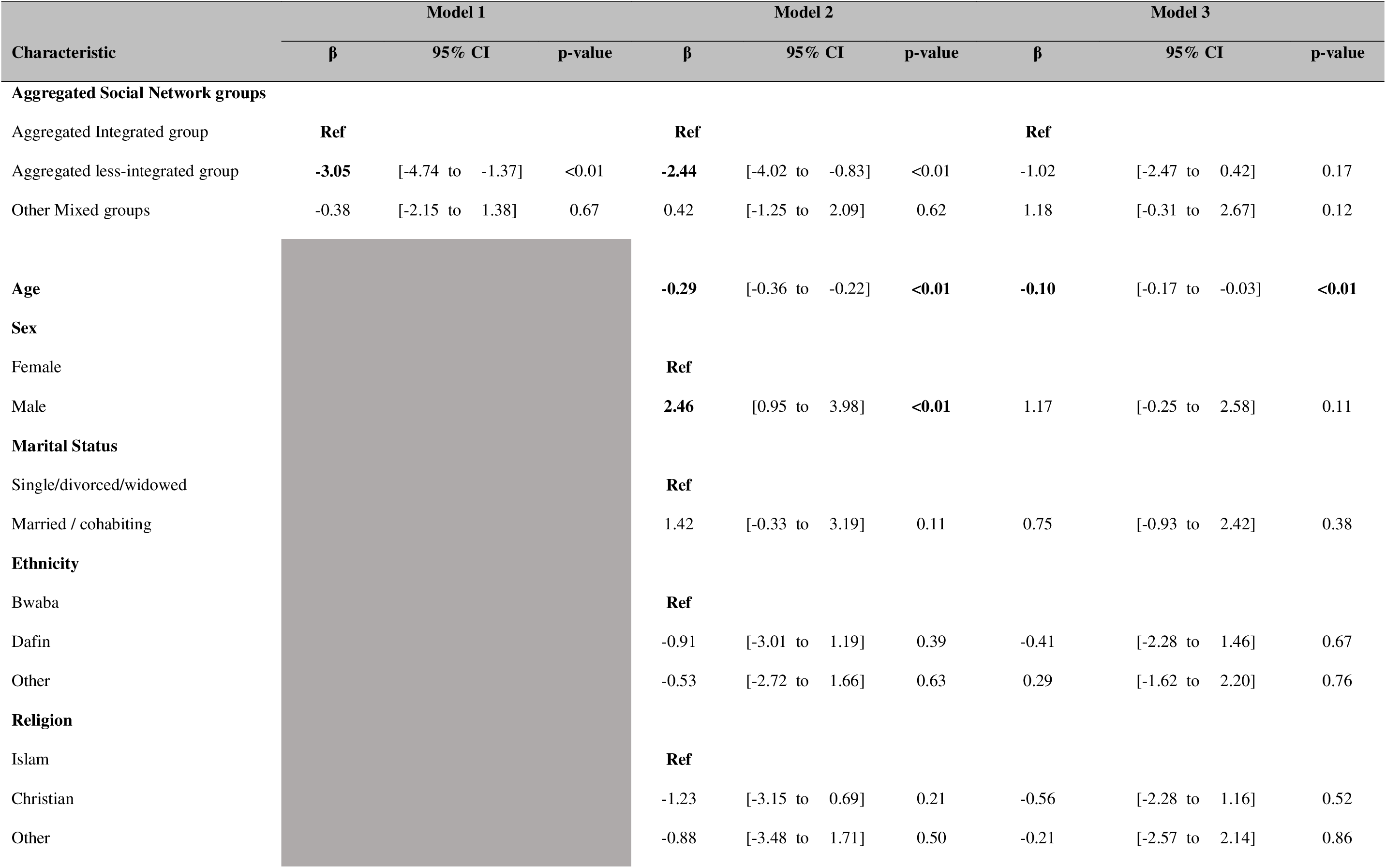
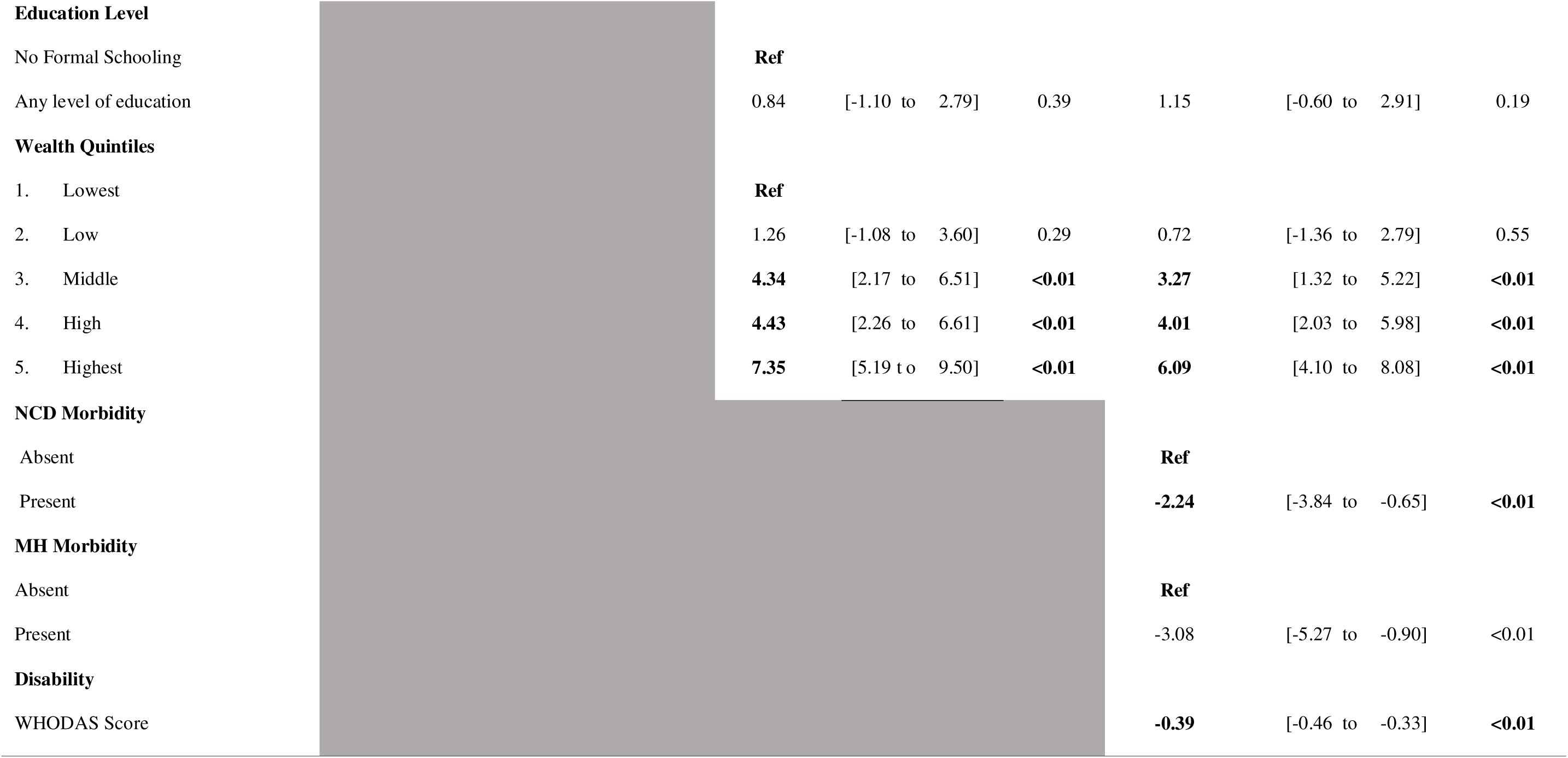
Weighted linear regression models for relationship between WHO quality of life score and Aggregated social network types, in the presence of sociodemographic and morbidity factors (n=2390)

## Discussion

This study describes the social networks of older adults in a rural setting of one of the lowest income countries in the world, and their association with the QoL. We found that the older adults of Nouna region of Burkina Faso have three common but distinct types of PANT social networks (Family Dependent, Locally Integrated, and Mixed) in near similar proportions. Having the less integrated social networks (Private Restricted or the Locally self-contained type) and the community oriented Wider Community-Focussed type were much less common in Nouna, compared to findings from other HICs and LMICs [2, 8, 13, 14]. Therefore, our attempted categorisation of the broader aggregated integrated and aggregated less-integrated groups appears to provide a more realistic representation of the social networks of these older adults in Nouna.

We found that having the least common PANT network type, the Wider Community-Focussed social network, was significantly associated with a better QoL, although this association might have been exaggerated due to very low number of participants having this type of social network. Whereas in comparison to the Wider Community-Focussed or Locally Integrated social network types, having the less-integrated networks (Family Dependent or Private Restricted) were associated with a worse QoL. That the associations between both Private Restricted and aggregate less integrated network types and QoL diminished when controlling for sociodemographic and disappeared when controlling for morbidity variables, suggests that there may be an ability to modify this relationship. However, that many of the sociodemographic variables included (sex, age, and wealth) and morbidity factors require structural, fiscal, or cultural inputs to effect change, means that interventions are unlikely to have immediate benefits.

The distribution of the PANT social networks that we found appears to reflect Wegner’s hypothesis that older adults in communities with stable populations are more likely to have Locally Integrated networks, and that older persons in stable rural settings are likely to have Locally Integrated or Family Dependent networks [5]. Studies done in MICs in rural settings have also, largely shown that having a Locally Integrated group is most frequent [8]. In HICs, the distribution of PANT social network types among older adults varies based on the age of study populations and the study settings. For example, many rural communities in the United Kingdom have in-migration of economically wealthy retired older adults, and these communities tend to have a good representation of Wider Community-Focussed social network types [40]. Whilst some other studies conducted in less wealthy stable communities report that older persons have more restricted or family oriented social network types [13, 14, 41].

Males were more likely to have community oriented social networks (Locally Integrated and Wider Community-Focussed networks) in Nouna whereas females predominantly had less integrated social networks (Locally Self-Contained or Private Restricted). Males and females were roughly equal in having a Family Dependent social network. Similar sex-related differences have been reported from some MICs such as rural India and the Dominican Republic, where older women were more likely to have less community-integrated social networks [8, 42]. Different traditions and cultural practices, as well as the effect of patriarchal societal structures in some settings such as Nouna, could be reasons for sex differences in social networks [43, 44]. Indeed, such patriarchal societal practices are more prominent in the rural areas where poverty and illiteracy with lower rates of schooling are common among females, and its impact is maximally experienced by elderly, single or widowed females [43, 45]. Conversely, males in these societies have greater opportunities to widely integrate external to the family [43, 46, 47]. Younger age, better income and wealth status, and absence of disease and disability are proven factors associated with a better QoL [6].

Our study has several limitations. The high rate (22.4%) of non-response to the PANT questions was an important limitation in our study. This had been attributed to a misinterpretation of a skipping step in the CHAS questionnaire by some of the data collectors, whereby they had mistakenly skipped the section with PANT questions. This resulted in the loss of an important opportunity to collected additional data that could have yielded stronger results. However, a comparative analysis of the sociodemographic and morbidity characteristics of this non-response group was found to be similar to the responded group and hence, the effect this high non-response rate on the study findings was expected to be minimal. Choosing an appropriate tool for analysing and categorising community members into the social network groups is important, and multiple questionnaires exist for this purpose [48, 49]. Nevertheless, the PANT typology has been successfully validated in multiple other countries including in HICs and MICs [8, 50], and we tested the questionnaires with data collectors to ensure local relevance and utility prior to administration. However, in analysing our results, it appears that for Nouna, simply using the integrated and less-integrated aggregated network groups might provide a better overview of the social network types than when using the PANT typology. The use of self-reported data on NCDs could have led to an under or over estimation of the status of NCD morbidity, but it is unclear if this misreporting would be strongly associated with the exposure or outcome. In addition, although our study was conducted in a mostly rural region of Burkina Faso and the findings might be similar to most rural parts of the country, it might not reflect a true distribution of the social networks in the more affluent, urban areas of Burkina Faso. However, given the similarity of our results with those reported from other rural MIC settings, and considering the socio-economic similarities of the rural regions of low-income and middle-income settings, we understand that our results might be generalisable to other similar low-income settings in other parts of the world [8, 51].

In summary, this study highlights that in this rural low-income settings, the perceived QoL of older adults less influenced by their own social networks and are more impacted by the morbidity among the older populations. However, when considering the social networks alone, having a more community integrated network appears to be associated with a better QoL. Since social networks cannot be built within a short period, but are created and evolve throughout the lifetime, developing strong integrated social support networks needs to be encouraged when persons are young and active. This requires socio-cultural and economic change. In the immediate time, however, our results combine with those of others to highlight that isolated persons in rural communities in LMICs may have poor quality of life and other health outcomes. Awareness of these associations could result in interventions to improve quality of life and health outcomes that go beyond changes in social network structure. Furthermore, it is essential that more research is needed to further identify the social networks patterns among the older adults in other low-income settings, and how it affects their QoL, which will enable the provision of better care for elderly in these low-resourced settings.

## Author contributions

Conceptualization: JD, AS, GH, CG, MI, LH, MDW, SA-B, MB, BC, LO

Data curation: DG-P, IW, GH, SA-B

Formal analysis: IW, DG-P, GH, JD

Funding acquisition: JD, AS

Investigation: JD, AS, LO, BC, MB, GH

Methodology: JD, AS, GH, MDW, LH, CG, MI, LO, BC, MB

Project administration: JD, AS, LO, BC, MB

Writing – original draft: IW, DG-P

Writing – review & editing: DGP, GH, SA-B, CG, MI, LH, LO, BC, MB, AS, JD

## Data availability statement

De-identified individual participant data and analysis code will be made available upon reasonable request to the corresponding author.

## Ethics approval and consent to participate

Ethics approval was obtained from the Ethics committee of the Ministry of Health, Burkina Faso (2018-5-053).

## Consent for publication

All authors had full access to all the data in the study and accept final responsibility for the decision to submit for publication.

## Acknowledgement

All study participants from the Nouna HDSS region, and the CRSN study team from Nouna, Burkina Faso.

