## Supplementary material for "Social networks and their association with quality of life among older adults in rural Burkina Faso"

#### **Supplementary Material 1:** Conceptual framework of factors affecting Quality of Life (QoL)

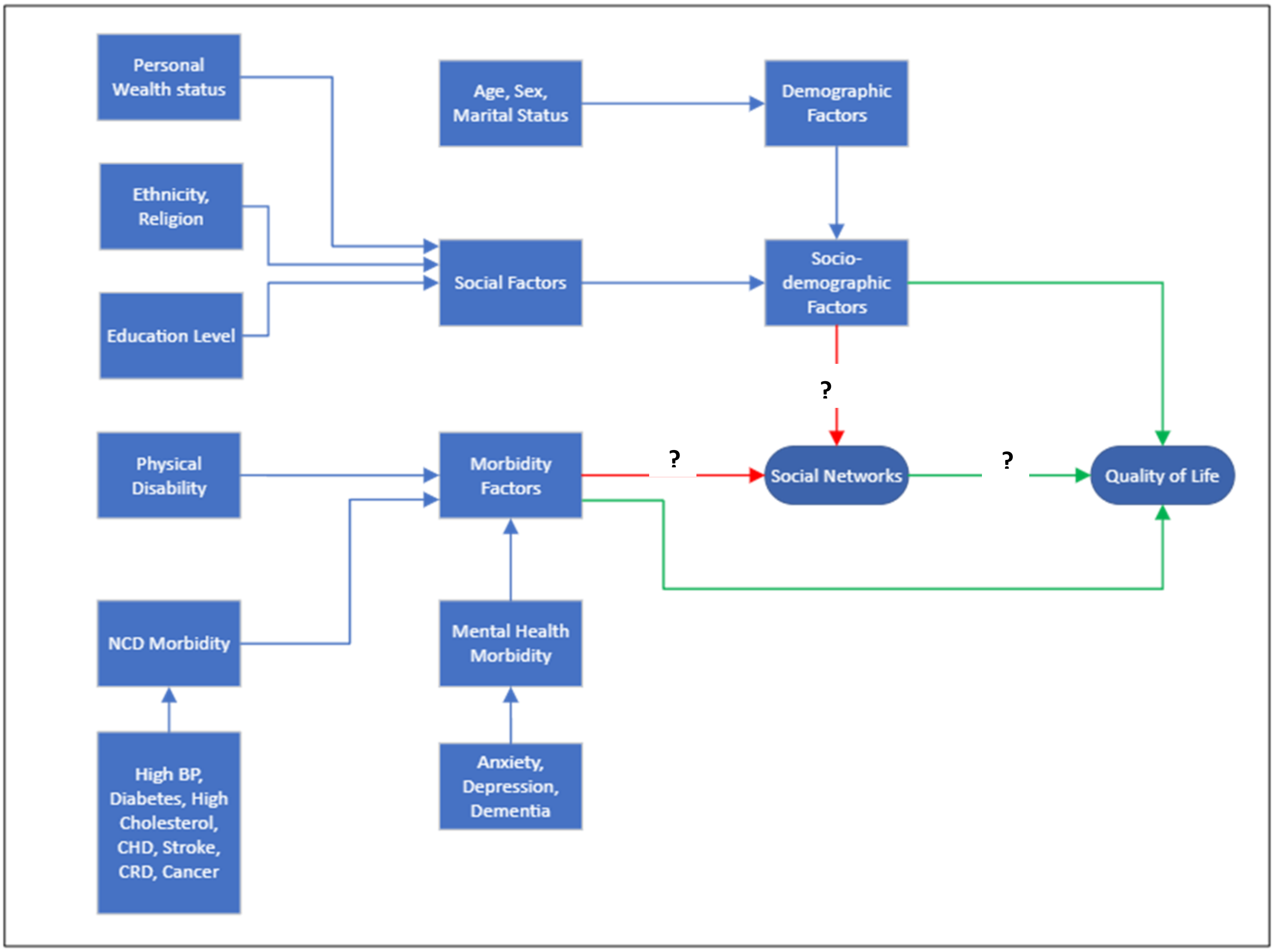

*Figure 1 - Conceptual Framework on factors affecting the Quality of Life, among older adults and the status of Social Networks,*

**List of publications used towards developing the Conceptual framework on factors affecting quality of life and the status of social networks:**

### **Supplementary Material 2:** Assessment of the PANT Social Network types

**Algorithm for Assessing the Social Network typology** [11]:

For each question, the relevant response code is identified. The relevant social network type for that question is identified in the subsequent columns. (E.g. If the response for Q.1 was 1 – 5 miles", the response code number is 2, and the relevant social network is LI. Once all 8 questions are completed, the total number of marks for each of the 5 Social Network typology columns are summed and the social network with the greatest number of marks s that to which the participant belongs. Where two or more networks receive equally high scores, the social network type is determined as mixed.

*Wenger's PANT Social Network Typology Assessment Instrument [11]*

| Question | Code | | FD | LI | LSC | WCF | PR |
|---|---|---|---|---|---|---|---|
| 1. How far away, in distance does your nearest child or other relative live? Do not include spouse | No relatives<br>Same house/ within 1 mile<br>1–5 miles<br>6–15 miles<br>16–50 miles<br>50+ miles | 0<br>1<br>2<br>3<br>4<br>5 | 1 | 2,3 | 3,4 | 4,5 | 4,5 |
| 2. If you have any children, where does your nearest child live? | No children<br>Same house/ within 1 mile<br>1–5 miles<br>6–15 miles<br>16–50 miles<br>50+ miles | 0<br>1<br>2<br>3<br>4<br>5 | 1,2 | 1,2,3 | 0,3,4 | 5 | 5 |
| 3. If you have any living sisters or brothers, where does your nearest sister or brother live? | No sister/brother<br>Same house/ within 1 mile<br>1–5 miles<br>6–15 miles<br>16–50 miles<br>50+ miles | 0<br>1<br>2<br>3<br>4<br>5 | 1,2 | 1,2,3 | 2,3,4 | 0,5 | 0,5 |
| 4. How often do you see any of your children or other relatives to speak to? | Never/no relative<br>Daily<br>2-3 times a week<br>At least weekly<br>At least monthly<br>Less often | 0<br>1<br>2<br>3<br>4<br>5 | 1,2 | 1,2 | 3,4 | 4,5 | 0,5 |
| 5. If you have friends in this community/neighbourhood, how often do you have a chat or do something with one of your friends? | Never/no friends<br>Daily<br>2-3 times a week<br>At least weekly<br>At least monthly<br>Less often | 0<br>1<br>2<br>3<br>4<br>5 | 4,5 | 1,2,3 | 4,5 | 2,3 | 0,5 |
| 6. How often do you see any of your neighbours to have a chat with or do something with? | Never/no neighbour contacts<br>Daily<br>2-3 times a week | 0<br>1<br>2 | 0,4,5 | 1,2,3 | 3,4 | 3,4 | 0,5 |

| | | | | | | | |
|---|---|---|---|---|---|---|---|
| | At least weekly<br>At least monthly<br>Less often | 3<br>4<br>5 | | | | | |
| 7. Do you attend any religious meetings? | Yes, regularly (at least once a month)<br>Yes, occasionally<br>No | 1<br>2<br>0 | 2 | 1 | 0,2 | 1,2 | 0 |
| 8. Do you attend meetings of any community/ neighbourhood or social groups, such as old people's clubs, lectures or anything like that? | Yes, regularly (at least once a month)<br>Yes, occasionally<br>No | 1<br>2<br>0 | 0,2 | 1 | 0,2 | 1 | 0 |
| **Network Type (Highest Number)** | | | | | | | |

#### **Supplementary Material 3:** Mixed group in the PANT social network analysis

*Table 1 – Analysis of Mixed group of Social Network types*

| | **Total (n=730)** | | **Women (n=450)** | | **Men (n=280)** | |
|---|---|---|---|---|---|---|
| | **N** | **%** | **N** | **%** | **N** | **%** |
| **Mixed group –** (LI & WCF characteristics) | 7 | 0.96% | 6 | 1.3% | 1 | 0.0 |
| **Mixed group –** (mixture of FD, PR and LSC characteristics) | 56 | 7.67% | 42 | 9.3% | 14 | 0.1 |
| **Other Mixed groups:** | | | | | | |
| LI & FD characteristics | 618 | 84.7% | 372 | 82.7% | 246 | 0.9 |
| LI & PR characteristics | 7 | 1.0% | 7 | 1.6% | - | - |
| LI & LSC characteristics | 18 | 2.5% | 13 | 2.9% | 5 | 0.0 |
| WCF & FD characteristics | 1 | 0.1% | 1 | 0.2% | - | - |
| WCF & PR characteristics | 4 | 0.5% | - | - | 4 | 0.0 |
| WCF & LSC characteristics | 1 | 0.1% | - | - | 1 | 0.0 |
| Multiple type characteristics | 18 | 2.5% | 9 | 2.0% | 9 | 0.0 |

**Supplementary Material 4:** Sociodemographic and morbidity factors related to each Social Network type inclusive of Mixed type (n=2390)

| Characteristic | Total | Unweighted % | Weighted % | Social Network Types (No. and Weighted %) *Except where indicated, shows Median and Inter Quartile Range (IQR)* | | | | | | | | | | | |
|---|---|---|---|---|---|---|---|---|---|---|---|---|---|---|---|
| | | | | LI | % | FD | % | LSC | % | PR | % | WCF | % | Mixed | % |
| **Total** | **2390** | | | **845** | **35.36%** | **725** | **30.33%** | **29** | **1.21%** | **51** | **2.13%** | **10** | **0.42%** | **730** | **30.54%** |
| **Age (in years)** | | | | | | | | | | | | | | | |
| Median | 55 | | | 54 | | 54 | | 59 | | 69 | | 51 | | 55 | |
| IQR | 47 to 64 | | | 47 to 62 | | 47 to 64 | | 54 to 68 | | 56 to 79 | | 46 to 52 | | 47 to 65 | |
| **Sex** | | | | | | | | | | | | | | | |
| Male | 1056 | 44.2% | 44.2% | 430 | 47.6% | 317 | 43.7% | 10 | 28.3% | 14 | 18.2% | 5 | 59.6% | 280 | 37.1% |
| Female | 1334 | 55.8% | 55.8% | 415 | 52.4% | 408 | 56.3% | 19 | 71.7% | 37 | 81.8% | 5 | 40.3% | 450 | 62.9% |
| **Marital Status** | | | | | | | | | | | | | | | |
| Married or cohabiting | 1664 | 69.6% | 62.5% | 626 | 69.7% | 519 | 63.6% | 14 | 33.5% | 23 | 35.1% | 8 | 82.9% | 474 | 55.6% |
| Single, divorced or widowed | 723 | 30.2% | 37.5% | 217 | 30.3% | 206 | 36.3% | 15 | 66.5% | 28 | 64.9% | 2 | 17.1% | 255 | 44.4% |
| **Ethnicity** | | | | | | | | | | | | | | | |
| Dafin | 772 | 32.3% | 31.4% | 272 | 30.4% | 234 | 33.6% | 7 | 24.4% | 9 | 17.2% | 6 | 54.1% | 244 | 31.4% |
| Bwaba | 801 | 33.5% | 32.6% | 326 | 39.5% | 231 | 29.4% | 12 | 38.0% | 15 | 28.2% | 2 | 15.1% | 215 | 27.6% |
| Other | 817 | 34.2% | 36.0% | 247 | 30.1% | 260 | 36.9% | 10 | 37.5% | 27 | 54.6% | 2 | 30.7% | 271 | 41.0% |
| **Religion** | | | | | | | | | | | | | | | |
| Muslim | 1228 | 51.4% | 51.3% | 371 | 42.8% | 401 | 56.5% | 12 | 40.6% | 28 | 54.2% | 5 | 57.4% | 411 | 51.3% |
| Christian | 843 | 35.3% | 34.3% | 335 | 39.2% | 227 | 29.7% | 12 | 37.7% | 19 | 36.6% | 4 | 27.5% | 246 | 34.3% |
| Other | 319 | 13.3% | 14.4% | 139 | 18.0% | 97 | 13.9% | 5 | 21.7% | 4 | 9.2% | 1 | 15.1% | 73 | 14.4% |
| **Education Level** | | | | | | | | | | | | | | | |
| No Formal Schooling | 1982 | 82.9% | 83.4% | 689 | 82.1% | 609 | 83.3% | 21 | 81.2% | 44 | 91.8% | 7 | 71.1% | 612 | 84.7% |
| Any level of education | 408 | 17.1% | 16.6% | 156 | 17.9% | 116 | 16.7% | 8 | 18.8% | 7 | 8.1% | 3 | 28.9% | 118 | 15.3% |

| Characteristic | Total | Unweighted % | Weighted % | **Social Network Types (No. and Weighted %)** *Except where indicated, shows Median and Inter Quartile Range (IQR)* | | | | | | | | | | | |
|---|---|---|---|---|---|---|---|---|---|---|---|---|---|---|---|
| | | | | **LI** | **%** | **FD** | **%** | **LSC** | **%** | **PR** | **%** | **WCF** | **%** | **Mixed** | **%** |
| **Wealth Quintiles** | | | | | | | | | | | | | | | |
| 1 - Lowest | 411 | 17.2% | 18.4% | 154 | 20.4% | 111 | 14.9% | 3 | 8.9% | 13 | 31.8% | 3 | 27.1% | 127 | 18.4% |
| 2 - Low | 418 | 17.5% | 17.4% | 152 | 18.8% | 102 | 12.9% | 6 | 20.8% | 8 | 12.9% | 1 | 0.0% | 149 | 17.4% |
| 3 - Middle | 442 | 18.5% | 16.8% | 175 | 17.4% | 139 | 19.2% | 4 | 13.7% | 7 | 21.9% | 1 | 0.0% | 116 | 16.8% |
| 4 - High | 516 | 21.6% | 20.8% | 173 | 19.7% | 172 | 22.8% | 9 | 35.9% | 8 | 9.5% | 2 | 30.5% | 1523 | 20.8% |
| 5 - Highest | 603 | 25.2% | 26.5% | 191 | 23.7% | 201 | 30.1% | 7 | 20.7% | 15 | 23.8% | 3 | 42.4% | 186 | 26.5% |
| **Non-Communicable Disease (NCD) Morbidity** | | | | | | | | | | | | | | | |
| Absent | 1401 | 58.6% | 55.6% | 514 | 58.1% | 408 | 52.8% | 19 | 68.6% | 32 | 59.2% | 4 | 40.9% | 424 | 55.6% |
| Present | 989 | 41.4% | 44.4% | 331 | 41.5% | 317 | 47.2% | 10 | 31.4% | 19 | 40.8% | 6 | 59.1% | 306 | 44.4% |
| **Anxiety** | | | | | | | | | | | | | | | |
| Absent | 2166 | 90.6% | 89.6% | 791 | 92.9% | 631 | 85.5% | 24 | 85.8% | 43 | 79.4% | 9 | 100.0% | 668 | 90.3% |
| Present | 224 | 9.4% | 10.4% | 54 | 7.1% | 94 | 14.5% | 5 | 14.2% | 8 | 20.6% | 1 | 0.0% | 62 | 9.7% |
| **Depression** | | | | | | | | | | | | | | | |
| Absent | 2163 | 90.5% | 89.3% | 793 | 92.9% | 633 | 85.8% | 24 | 77.8% | 44 | 82.5% | 10 | 100.0% | 659 | 89.5% |
| Present | 227 | 9.5% | 10.6% | 52 | 7.13% | 92 | 14.2% | 5 | 22.2% | 7 | 17.5% | 0 | 0.0% | 71 | 10.5% |
| Total | 2390 | | | 845 | 35.4% | 725 | 30.3% | 29 | 1.2% | 51 | 2.1% | 10 | 0.4% | 730 | 30.5% |
| **Dementia** | | | | | | | | | | | | | | | |
| Absent | 2305 | 96.4% | 95.1% | 825 | 96.9% | 704 | 96.6% | 26 | 83.3% | 43 | 74.6% | 10 | 100.0% | 697 | 93.7% |
| Present | 85 | 3.6% | 4.8% | 20 | 3.1% | 21 | 3.4% | 3 | 16.7% | 8 | 25.4% | 0 | 0.0% | 33 | 6.3% |
| **Overall Mental Health Morbidity*** | | | | | | | | | | | | | | | |
| Absent | 2040 | 85.4% | 82.9% | 761 | 88.2% | 594 | 79.3% | 21 | 67.9% | 36 | 62.1% | 9 | 100.0% | 619 | 82.9% |
| Present | 350 | 14.6% | 17.0% | 84 | 11.8% | 131 | 20.7% | 8 | 32.0% | 15 | 37.9% | 1 | 0.0% | 111 | 17.0% |
| **Disability (normalised WHO DAS 10.0 score)** | | | | | | | | | | | | | | | |
| Median | 18.7 | | | 18.7 | | 18.7 | | 22.9 | | 27.1 | | 18.7 | | 18.7 | |
| IQR | 18.7 to 25.0 | | | 18.7 to 22.9 | | 18.7 to 27.1 | | 18.75 to 35.4 | | 18.7 to 52.1 | | 18.7 to 22.9 | | 18.7 to 27.1 | |
| **Quality of Life (normalised WHOQOL score)** | | | | | | | | | | | | | | | |
| Median | 59.4 | | | 62.5 | | 59.4 | | 59.4 | | 53.1 | | 65.6 | | 59.4 | |
| IQR | 50.0 to 65.6 | | | 50.0 to 65.6 | | 46.9 to 65.6 | | 46.9 to 65.6 | | 40.6 to 62.5 | | 65.6 to 68.7 | | 50.0 to 65.6 | |

*Anxiety, Depression and Dementia combined

#### **Supplementary Material 5:** Weighted multinomial logistic regression model of PANT social network types inclusive of Mixed type (n=2390)

*Table 2 – Weighted multinomial logistic regression model of PANT social network types and their association with the sociodemographic and morbidity factors, in reference to Locally Integrated network type*

| Characteristic | Family Dependent | | | Private Restricted | | | PANT Social Network types<br>Locally Self-Contained | | | Wider Community-Focused | | | Mixed type | | |
|---|---|---|---|---|---|---|---|---|---|---|---|---|---|---|---|
| | **RRR** | **95% CI** | **p-value** | **RRR** | **95% CI** | **p-value** | **RRR** | **95% CI** | **p-value** | **RRR** | **95% CI** | **p-value** | **RRR** | **95% CI** | **p-value** |
| **Age** | | | | | | | | | | | | | | | |
| Age in years | 0.99 | 0.98-1.01 | 0.43 | **1.07** | 1.02-1.13 | 0.00 | **1.06** | 1.01-1.12 | 0.02 | **0.92** | 0.86-0.98 | 0.01 | 0.99 | 0.98-1.01 | 0.99 |
| **Sex** | | | | | | | | | | | | | | | |
| Female | Ref | | | Ref | | | Ref | | | Ref | | | Ref | | |
| Male | 0.79 | 0.58-1.08 | 0.14 | **0.22** | 0.06-0.71 | 0.01 | 0.52 | 0.17-1.64 | 0.27 | 1.12 | 0.15-8.42 | 0.91 | 0.64 | 0.47-0.87 | 0.05 |
| **Marital Status** | | | | | | | | | | | | | | | |
| Single/ divorced/ widowed | Ref | | | Ref | | | Ref | | | Ref | | | Ref | | |
| Married / cohabiting | 0.85 | 0.60-1.22 | 0.40 | 0.95 | 0.31-2.87 | 0.94 | 0.48 | 0.16-1.40 | 0.18 | 1.01 | 0.03-30.2 | 0.99 | **0.69** | 0.48-0.99 | 0.03 |
| **Ethnicity** | | | | | | | | | | | | | | | |
| Bwaba | Ref | | | Ref | | | Ref | | | Ref | | | Ref | | |
| Dafin | 1.00 | 0.66-1.53 | 0.99 | 0.51 | 0.14-1.82 | 0.30 | 0.80 | 0.19-3.37 | 0.76 | 6.19 | 0.70-54.5 | 0.10 | 1.05 | 0.68-1.62 | 0.81 |
| Other | 1.05 | 0.68-1.64 | 0.81 | 2.32 | 0.74-7.25 | 0.15 | 1.28 | 0.29-5.54 | 0.74 | 3.68 | 0.21-63.1 | 0.37 | 1.44 | 0.92-2.24 | 0.11 |
| **Religion** | | | | | | | | | | | | | | | |
| Islam | Ref | | | Ref | | | Ref | | | Ref | | | Ref | | |
| Christian | **0.61** | 0.42-0.88 | 0.01 | 0.72 | 0.25-2.04 | 0.54 | 1.11 | 0.32-3.83 | 0.87 | 1.02 | 0.13-7.97 | 0.98 | **0.66** | 0.45-0.96 | 0.03 |
| Other | 0.62 | 0.37-1.04 | 0.07 | 0.28 | 0.06-1.34 | 0.11 | 1.22 | 0.23-6.43 | 0.81 | 1.89 | 0.26-13.5 | 0.53 | **0.47** | 0.27-0.82 | 0.01 |
| **Education Level** | | | | | | | | | | | | | | | |
| No Formal Schooling | Ref | | | Ref | | | Ref | | | Ref | | | Ref | | |
| Any level of education | 1.01 | 0.68-1.51 | 0.93 | 1.45 | 0.34-6.28 | 0.61 | 2.36 | 0.76-7.30 | 0.14 | 1.34 | 0.39-4.61 | 0.65 | 1.15 | 0.76-1.73 | 0.49 |
| **Wealth Quintiles** | | | | | | | | | | | | | | | |
| 1. Lowest | Ref | | | Ref | | | Ref | | | Ref | | | Ref | | |
| 2. Low | 0.90 | 0.55-1.41 | 0.67 | 0.35 | 0.09-1.35 | 0.13 | 2.90 | 0.53-15.8 | 0.21 | NR | NR | NR | 1.11 | 0.71-1.72 | 0.63 |
| 3. Middle | 1.48 | 0.93-2.31 | 0.09 | 0.70 | 0.24-2.04 | 0.52 | 2.30 | 0.35-14.9 | 0.38 | NR | NR | NR | 0.82 | 0.51-1.31 | 0.41 |
| 4. High | 1.44 | 0.89-2.21 | 0.12 | **0.20** | 0.04-0.96 | 0.04 | 4.99 | 0.99-25.2 | 0.05 | 0.76 | 0.10-5.76 | 0.79 | 0.96 | 0.62-1.51 | 0.89 |
| 5. Highest | 1.57 | 0.98-2.42 | 0.05 | 0.43 | 0.14-1.27 | 0.13 | 2.58 | 0.43-15.2 | 0.29 | 0.71 | 0.08-5.91 | 0.75 | 1.03 | 0.66-1.61 | 0.88 |
| **NCD Morbidity** | | | | | | | | | | | | | | | |
| Absent | Ref | | | Ref | | | Ref | | | Ref | | | Ref | | |

| | | | | | | | | | | | | | | | |
|---|---|---|---|---|---|---|---|---|---|---|---|---|---|---|---|
| Present | 1.28 | 0.92-1.79 | 0.14 | 1.18 | 0.52-2.68 | 0.70 | 0.64 | 0.21-1.91 | 0.42 | 2.00 | 0.36-10.9 | 0.43 | 1.04 | 0.75-1.46 | 0.77 |
| **MH Morbidity** | | | | | | | | | | | | | | | |
| Absent | Ref | | | Ref | | | Ref | | | Ref | | | Ref | | |
| Present | **1.67** | 1.11-2.51 | 0.01 | 2.15 | 0.82-5.65 | 0.12 | 2.14 | 0.84-5.44 | 0.11 | NR | NR | NR | 1.24 | 0.82-1.88 | 0.32 |
| **Disability** | | | | | | | | | | | | | | | |
| WHODAS Score | 1.01 | 0.99-1.02 | 0.06 | 1.07 | 0.99-1.04 | 0.20 | 1.01 | 0.97-1.03 | 0.74 | 0.99 | 0.93-1.06 | 0.93 | 1.01 | 1.00-1.03 | 0.71 |

RRR - Relative Risk ratio, 95% CI - 95% Confidence Interval, p-value - Probability value, Ref – Reference variable class, Significant associations are depicted with a bold font RRR.

#### **Supplementary Material 6:** Weighted linear regression models for relationship between WHO quality of life score and PANT social network types inclusive of the mixed type, in the presence of sociodemographic and morbidity covariates (n=2390)

*Table 3 - Weighted linear regression models for relationship between WHO quality of life score and social network typologies, in the presence of sociodemographic and morbidity covariates*

| | Model 1 | | | Model 2 | | | Model 3 | | |
|---|---|---|---|---|---|---|---|---|---|
| **Characteristic** | **β** | **95% CI** | **p-value** | **β** | **95% CI** | **p-value** | **β** | **95% CI** | **p-value** |
| **Social Network Typologies** | | | | | | | | | |
| Locally Integrated (LI) | **Ref** | | | **Ref** | | | **Ref** | | |
| Family Dependent (FD) | **-2.53** | [-4.31 to -0.75] | <0.01 | **-2.65** | [-4.32 to -0.99] | 0.02 | -1.41 | [-2.94 to 0.11] | 0.07 |
| Private Restricted (PR) | **-5.71** | [-11.13 to -0.29] | 0.04 | -0.25 | [-5.12 to 4.61] | 0.92 | 2.47 | [-2.44 to 7.38] | 0.32 |
| Locally Self-contained (LSC) | -1.94 | [-7.02 to 3.14] | 0.45 | 2.05 | [-2.02 to 6.35] | 0.35 | 3.01 | [-1.65 to 7.89] | 0.21 |
| Wider Community-Focussed (WCF) | **11.88** | [6.58 to 17.17] | <0.01 | **8.60** | [3.19 to 14.00] | <0.01 | **8.69** | [4.10 to 13.27] | <0.01 |
| Mixed type of Social Network | -0.38 | [-2.13 to 1.36] | 0.67 | 0.64 | [-1.03 to 2.28] | 0.45 | **1.53** | [0.06 to 3.00] | 0.04 |
| **Age** | | | | **-0.31** | [-0.37 to -0.22] | <0.01 | **-0.11** | [-0.18 to -0.04] | 0.00 |
| **Sex** | | | | | | | | | |
| Female | | | | Ref | | | | | |
| Male | | | | **2.58** | [1.08 to 4.09] | <0.01 | 1.30 | [-0.10 to 2.71] | 0.07 |
| **Marital Status** | | | | | | | | | |
| Single/divorced/widowed | | | | Ref | | | | | |
| Married / cohabiting | | | | 1.45 | [-0.29 to 3.19] | 0.10 | 0.77 | [-0.86 to 2.41] | 0.35 |
| **Ethnicity** | | | | | | | | | |
| Bwaba | | | | Ref | | | | | |
| Dafin | | | | -0.96 | [-3.05 to 1.12] | 0.37 | -0.45 | [-2.30 to 1.41] | 0.64 |
| Other | | | | -0.69 | [-2.88 to 1.49] | 0.53 | -0.11 | [-1.79 to 2.01] | 0.91 |
| **Religion** | | | | | | | | | |
| Islam | | | | Ref | | | | | |
| Christian | | | | -1.24 | [-3.15 to 0.68] | 0.20 | 0.57 | [-2.28 to 1.14] | 0.51 |
| Other | | | | -0.96 | [-3.57 to 1.63] | 0.47 | -0.25 | [-2.59 to 2.09] | 0.83 |
| **Education Level** | | | | | | | | | |
| No Formal Schooling | | | | Ref | | | | | |
| Any level of education | | | | 0.76 | [-1.18 to 2.71] | 0.44 | 1.07 | [-0.68 to 2.80] | 0.23 |

| | | | | | | | |
|---|---|---|---|---|---|---|---|
| **Wealth Quintiles** | | | | | | | |
| 1. Lowest | | Ref | | | | | |
| 2. Low | | 1.29 | [-1.06 to 3.64] | 0.28 | 0.68 | [-1.04 to 2.77] | 0.52 |
| 3. Middle | | **4.47** | [2.29 to 6.57] | <0.01 | **3.39** | [1.42 to 5.36] | <0.01 |
| 4. High | | **4.39** | [2.21 to 5.56] | <0.01 | **3.99** | [20.02 to 5.97] | <0.01 |
| 5. Highest | | **7.42** | [5.26 to 9.57] | <0.01 | **6.15** | [4.16 to 8.13] | <0.01 |
| **NCD Morbidity** | | | | | | | |
| Absent | | | | | Ref | | |
| Present | | | | | **-2.21** | [-3.80 to -0.62] | <0.01 |
| **MH Morbidity** | | | | | | | |
| Absent | | | | | Ref | | |
| Present | | | | | **-3.09** | [-5.26 to -0.91] | <0.01 |
| **Disability** | | | | | | | |
| WHODAS Score | | | | | **-0.38** | [-0.45 to -0.32] | <0.01 |

β – Linear Regression Coefficient, 95% CI – 95% Confidence Interval, p-value – Probability value, Ref – Reference variable class, Significant coefficients (β) are highlighted in bold font.
